# Impact of Iron Deficiency on HbA1c Accuracy in Monitoring Glycaemic Control in Non-Anaemic Type 2 Diabetes individuals: A prospective longitudinal study

**DOI:** 10.1101/2025.04.28.25326576

**Authors:** Rogers John Mukasa, Nathan Mubiru, Isaac Sekitoleko, Ronald Makanga, Hubert Nkabura, Terry Ongaria, Viola Mugamba, Wisdom Nakanga, Moffat J. Nyirenda, Anxious J. Niwaha

## Abstract

**Purpose:** Results from a few studies have been conflicting as to whether iron deficiency alters HbA1c reliability and the mechanisms on how iron might affect HbA1c reliability are not fully known. We aimed to compare the relationship between HbA1c and mean glucose concentrations measured by continuous glucose monitoring in iron replete and iron deplete states among non-anaemic type 2 diabetes mellitus (T2DM) patients.

**Methods:** We compared the differences in HbA1c between iron replete and deplete groups using the Chi-square test for categorical data and the Mann-Whitney U test for continuous data. We also evaluated the correlation between HbA1c and mean plasma glucose for both iron-replete and iron-deplete individuals using Pearson’s correlation and linear regression.

**Results:** A total of 146 of the 213 participants screened had complete data and were considered for final analysis. 43/146 (29.5%) had iron deficiency and 103 were iron replete. No significant difference was observed in HbA1c levels between iron-replete and iron-deplete individuals: 69 [51.0, 85.0] vs 62 [46.0, 83.0] mmol/mol, P 0.291). There was a strong positive correlation between HbA1c and mean plasma glucose concentration for both iron-replete and iron-deplete individuals (Pearson Correlation coefficient: 0.83 (0.76 – 0.91) and 0.93 (0.89 – 0.98), respectively).

**Conclusions:** HbA1c correlates well with mean blood glucose even in the iron deplete state amongst non-anaemic T2DM individuals. However, larger studies are needed to confirm these findings, particularly at screening and diagnostic thresholds.

## Introduction

Diabetes mellitus (DM) is a significant global health problem, currently affecting approximately 463 million individuals globally(1). Of these 336 million (approximately 4/5 of the world’s diabetic population) reside in low and middle income countries (LMIC)(2). Effective diabetes monitoring is essential for the appropriate titration of medication, enabling optimal glycaemic control and prevention of complications, thereby ensuring better health outcomes for individuals living with diabetes.

Glycated haemoglobin (HbA1c) is widely recognized as the gold standard for monitoring glycaemic control(3). However, it is increasingly evident that HbA1c can be influenced by non-glycaemic factors, potentially undermining its reliability as an accurate measure of long-term glycaemic exposure (4, 5). Iron deficiency (ID), the most common micronutrient deficiency worldwide(6) has been identified as one of the factors that may alter HbA1c levels. The proposed mechanisms underlying the interaction between ID and HbA1c remain poorly understood, but include; alteration of the haemoglobin globin chain quaternary structure in iron deficiency leading to rapid glycation, increased average erythrocyte life span in iron deficiency hence increased HbA1c and increased glycation of the proline terminal through peroxidation by the reduction of the anti-oxidant capacity of iron containing enzymes in the iron deficient state (7, 8).

Despite the existing literature on ID and HbA1c, significant gaps remain in our understanding of their interaction. Previous studies have methodological limitations, including small sample sizes and inadequate control for confounding factors such as anaemia and hemoglobinopathies. Consequently, the results are inconsistent and often fail to account for these confounders, leaving a critical gap in our understanding. For instance, while some studies suggest that HbA1c levels may be elevated in individuals with iron deficiency, others report reduced levels or no significant difference between the two groups(9).

Understanding the relationship between HbA1c and mean glucose levels in ID patients with type 2 diabetes is crucial particularly in LMICs, where both diabetes and iron deficiency are prevalent, and screening for ID among patients with diabetes is not a routine practice. Our study aimed to examine impact of ID on HbA1c as well as the relationship between HbA1c and mean glucose levels, as measured by continuous glucose monitoring (CGM), in both iron-deplete and iron-replete states among non-anaemic patients with type 2 diabetes participating in the OPTIMAL study.

## Study Methods

This was a prospective observational longitudinal study, conducted between 01^st^ August 2019 and 27^th^ February 2020, at two healthcare facilities: St. Francis Hospital Nsambya and Masaka Regional Referral Hospital in Uganda. The study enrolled type 2 diabetes mellitus patients, aged 18 years and above, who were three months post-diagnosis with no alterations in glucose-lowering therapy during the initial three months after screening. As shown in Fig. 1, we excluded pregnant women, critically ill patients, individuals requiring an immediate increase in glucose-lowering medication, those with anaemia, renal impairment, and haemoglobinopathies. All participants provided written consent prior to their involvement in the study.

**Fig 1.**
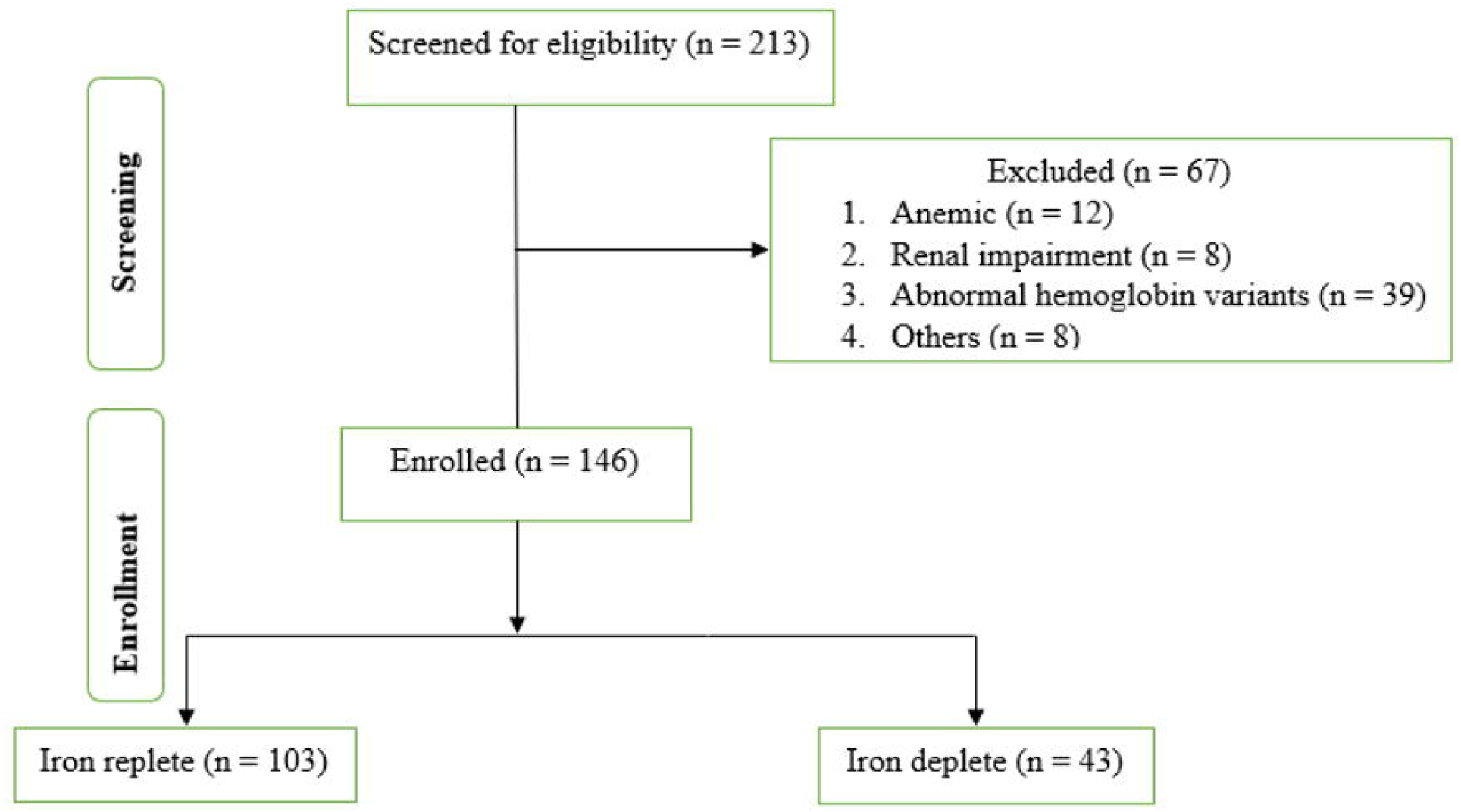
Participant flow chart.

Detailed description of the study procedures have been described elsewhere (10). In summary, participants attended three study visits. During Visit 1, they arrived in a non-fasted state, clinical and socio-demographic data, including age and sex, were collected using a standardized structured questionnaire. Trained research nurses obtained anthropometric measurements, recording weight in kilograms and height in meters. A non-fasting random blood sample (within 5 hours of a meal) was taken for laboratory analysis, which included assessments of Complete Blood Count (CBC), HbA1c (mmol/mol), glucose (mmol/L), and iron levels (μg/L). Continuous glucose monitoring was initiated using the Freestyle Libre Pro Flash Glucose Monitoring System (Abbott Laboratories, Illinois, USA), which continuously records interstitial glucose levels every 15 minutes for 14 days.

In Visit 2, participants returned in a fasted state (at least 8 hours after a meal) between days 7 and 10 of CGM for fasting glucose measurement. For Visit 3, held between days 12 and 14 from the baseline visit, participants returned in a non-fasted state (within 5 hours of a meal) for the downloading of CGM data and collection of a venous blood sample for HbA1c measurement.

### Laboratory analysis

Plasma glucose, HbA1c, renal function and full blood count were analysed at the MRC & LSHTM / UVRI clinical diagnostic laboratory services (CDLS). Serum ferritin was analysed at the Exeter clinical laboratory (Exeter, UK). HbA1c was measured with the Roche Cobas 6000 immunoassay while Hb, MCV, MCH and MCHC were measured by the Sysmex XN-1000™ analyser.

### Statistical analysis

Categorical data were presented using frequencies and percentages, while continuous data were presented as medians [Inter-quartile range (IQR)] due to the nonparametric nature of most data. We compared HbA1c levels across different iron categories based on serum ferritin levels — category 1 (<58μg/L); category 2 (58μg/L < 91μg/L); category 3 (91 < 138μg/L); category 4 (138 < 247μg/L); category 5 (≥ 247μg/L). We then subdivided the participants into two groups – iron replete and iron deplete using serum ferritin: iron deplete was defined as serum ferritin < 70 pmol/L and iron replete as serum ferritin ≥ 70 pmol/L.

Differences between groups were assessed using the Chi-square test for categorical data and the Mann-Whitney U test for continuous data. Pearson’s correlation was used to evaluate the correlation between HbA1c and mean plasma glucose for both iron-replete and iron-deplete individuals. Additionally, a sensitivity analysis was conducted by matching iron deplete (cases) and iron replete (controls) based on age and sex to control for potential confounding effects resulting from the initial large disparity in the case-to-control ratio. Pearson’s correlation was used to assess the correlation between HbA1c and mean plasma glucose within the matched groups, and the Mann-Whitney U test compared medians of HbA1c between the two matched groups. Linear regression analysis was used to assess effect of iron deficiency (exposure) on HbA1c (outcome) adjusting for apriori determined confounding factors i.e age, sex, duration of diabetes mellitus, history of hypertension and body mass index (BMI)(11-15). All statistical analyses were performed using Stata version 18.

### Ethical considerations

Ethical approval for the study was granted by the Research and Ethics committees of Uganda Virus Research Institute (UVRI-121/2019) and Uganda National Council of Science and Technology (UNCST) (HS 2588).

## Results

### Baseline and clinical characteristics of participants

A total of 146 participants were included in the study (Fig 1). Of these, 43 (29.5%) participants were classified as iron-deplete, while 103 (70.6%) were iron-replete. The prevalence of iron deficiency was higher in rural areas, with proportions of 22.9% among males and 42.2% among females. Additional data are given in (S1 graph). In urban areas, the proportions were lower, at 12.5% of males and 31.0% for females as shown in (S1 graph). No significant differences were observed between the iron-deplete and iron-replete groups concerning median age (55: IQR [50, 62] vs. 55 [45, 60], p = 0.323), BMI (27.3 [24.3, 30.2] vs. 27.1 [24.4, 30.7], p = 0.812), or duration of diabetes (Table 1).

**Table 1.**
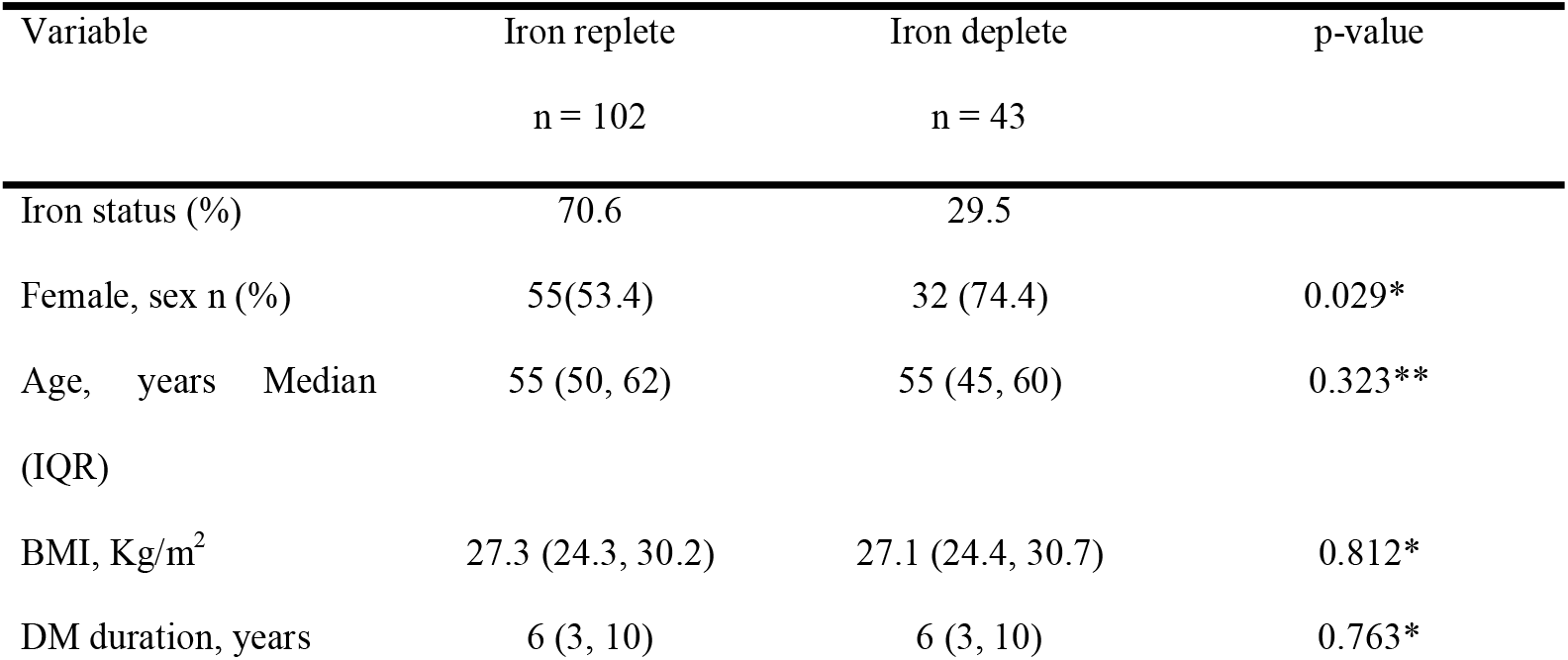
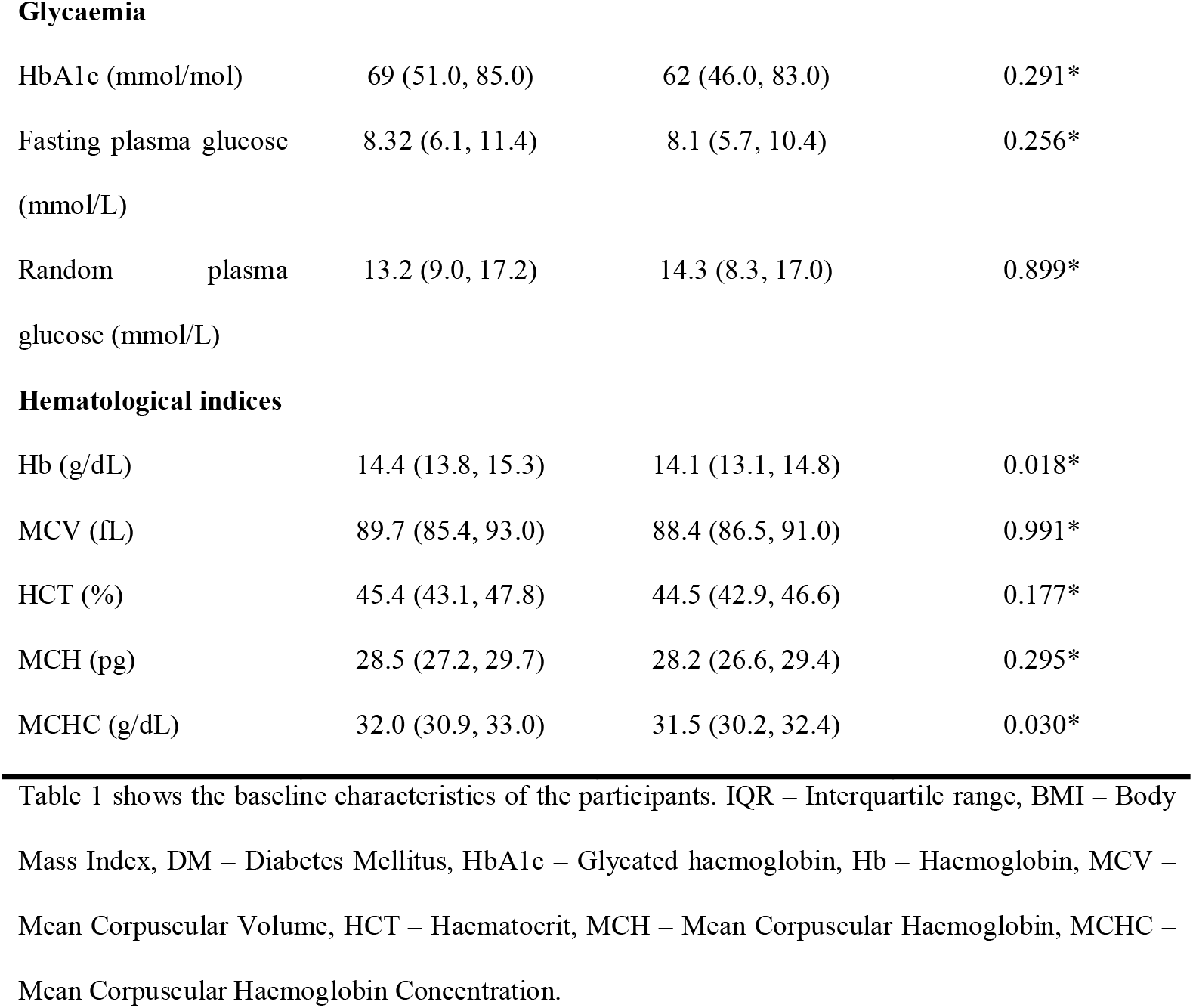
Baseline characteristics of the participants.

| Variable | Iron replete<br>n = 102 | Iron deplete<br>n = 43 | p-value |
| --- | --- | --- | --- |
| Iron status (%) | 70.6 | 29.5 |  |
| Female, sex n (%) | 55(53.4) | 32 (74.4) | 0.029* |
| Age, years Median<br>(IQR) | 55 (50, 62) | 55 (45, 60) | 0.323** |
| BMI, Kg/m<sup>2</sup> | 27.3 (24.3, 30.2) | 27.1 (24.4, 30.7) | 0.812* |
| DM duration, years | 6 (3, 10) | 6 (3, 10) | 0.763* |
| **Glycaemia** |  |  |  |
| HbA1c (mmol/mol) | 69 (51.0, 85.0) | 62 (46.0, 83.0) | 0.291* |
| Fasting plasma glucose<br>(mmol/L) | 8.32 (6.1, 11.4) | 8.1 (5.7, 10.4) | 0.256* |
| Random plasma<br>glucose (mmol/L) | 13.2 (9.0, 17.2) | 14.3 (8.3, 17.0) | 0.899* |
| **Hematological indices** |  |  |  |
| Hb (g/dL) | 14.4 (13.8, 15.3) | 14.1 (13.1, 14.8) | 0.018* |
| MCV (fL) | 89.7 (85.4, 93.0) | 88.4 (86.5, 91.0) | 0.991* |
| HCT (%) | 45.4 (43.1, 47.8) | 44.5 (42.9, 46.6) | 0.177* |
| MCH (pg) | 28.5 (27.2, 29.7) | 28.2 (26.6, 29.4) | 0.295* |
| MCHC (g/dL) | 32.0 (30.9, 33.0) | 31.5 (30.2, 32.4) | 0.030* |

Table 1 shows the baseline characteristics of the participants. IQR – Interquartile range, BMI – Body Mass Index, DM – Diabetes Mellitus, HbA1c – Glycated haemoglobin, Hb – Haemoglobin, MCV – Mean Corpuscular Volume, HCT – Haematocrit, MCH – Mean Corpuscular Haemoglobin, MCHC – Mean Corpuscular Haemoglobin Concentration.

### Erythrocytic indices but not glycaemic measures differed between iron-replete and iron-deplete individuals

The concentration of haemoglobin (Hb) was lower among iron-deficient participants compared to iron-replete individuals (14.4 [13.8, 15.3] vs 14.1 [13.1, 14.8] g/dl, P 0.018) (Table 1). The mean corpuscular haemoglobin concentration (MCHC) was also lower among iron-deficient participants compared to iron-replete individuals (32.0 [30.9, 33.0] vs 31.5 [30.2, 32.4] g/dl, P 0.030). However, no significant differences were found in other indices, including Mean Cell Volume (MCV), Mean Corpuscular Haemoglobin (MCH), and haematocrit (HCT), between the two groups.

In contrast, there was no difference HbA1c distribution by iron status (iron deplete (<70 μg/L) Vs iron replete (≥70 μg/L) groups) (Wilcoxon test: p = 0.291) (Fig 2). HbA1c levels were 69 [51.0, 85.0] vs 62 [46.0, 83.0] mmol/mol, P 0.291). Other measures of glycaemic burden did not differ significantly between iron-replete and iron-deplete individuals., fasting glucose (8.32 [6.1, 11.4] vs 8.1 [5.7, 10.4] mmol/L, P 0.256), and random glucose (13.2 [9.0, 17.2] vs 14.3 [8.3, 17.0] mmol/L, P 0.899). Furthermore, HbA1c levels did not differ significantly between the iron deplete cases and their matched iron replete controls (70.5 [55, 97] vs 64.0 [46.0, 80] mmol/mol, P 0.05).

**Fig 2.**
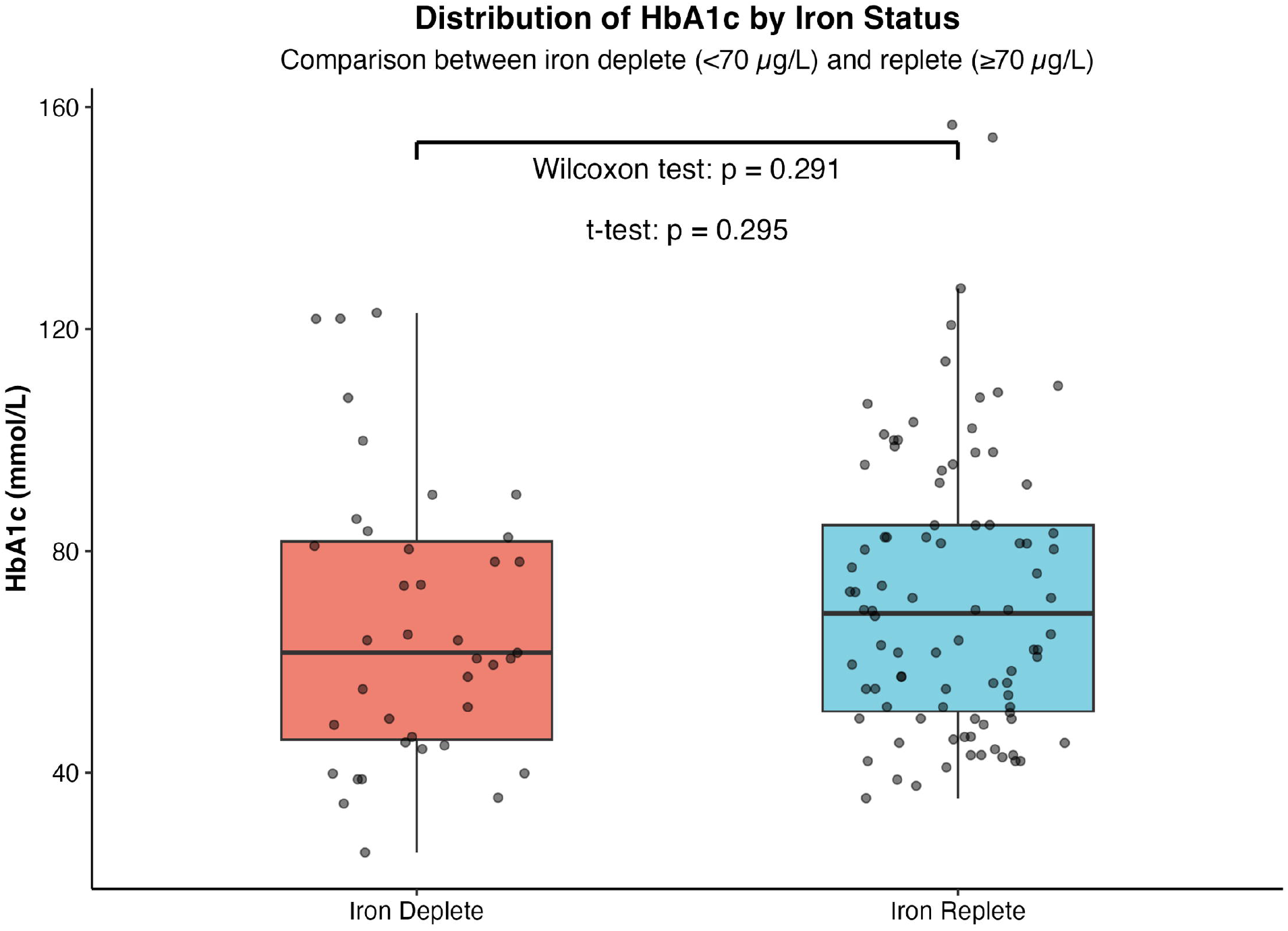
HbA1c distribution by iron by iron status. Distribution of HbA1c by iron status (iron deplete (<70 μg/L) Vs iron replete (≥70 μg/L) groups) (Wilcoxon test: p = 0.291). HbA1c distribution across five ferritin categories (<58, 58-90, 91-137, 138-246, ≥247 μg/L), showed an overlap between categories observed (Fig 3). Density distribution analysis further confirmed the substantial overlap between iron deplete and iron replete groups, with the iron replete group showing slightly higher density in the 80-120 mmol/L range (S2 graph).

**Fig 3.**
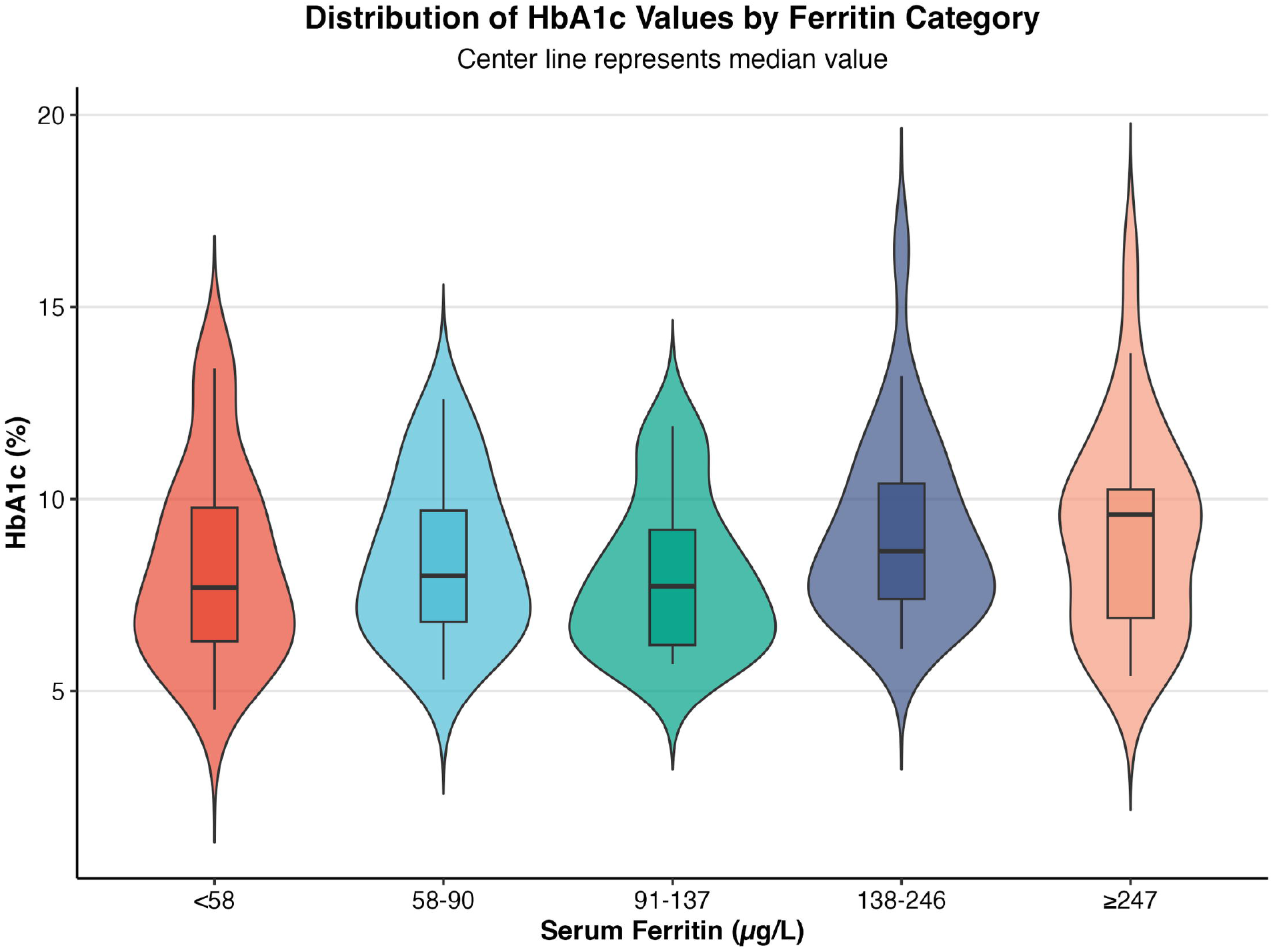
HbA1c distribution across iron categories. Mean of HbA1c across iron categories; serum ferritin levels — category 1 (<58μg/L); category 2 (58μg/L < 91μg/L); category 3 (91 < 138μg/L); category 4 (138 < 247μg/L); category 5 (≥ 247μg/L)

### Correlation between HbA1c with CGM glucose in both iron deplete and replete participants

Figures 4a and 4b illustrate the strong positive correlation that exists between HbA1c and mean blood glucose for both iron-replete and iron-deplete individuals respectively. Notably, the association was similar among iron-deplete participants compared to iron-replete individuals. A stronger correlation between HbA1c and mean plasma glucose levels was observed in the iron-deplete cases compared to the iron-replete controls (Pearson Correlation coefficient: 0.93 (0.88 – 0.98) and 0.88 (0.83 – 0.92), respectively). A strong positive correlation between HbA1c (mmol/mmol) and mean continuous glucose was also observed between both the iron-deplete cases (b) and their matched iron replete controls (a) as observed in S3 graph. (Pearson Correlation coefficient: 0.83 (0.76 – 0.91) and 0.93 (0.89 – 0.98), respectively).

**Fig 4.**
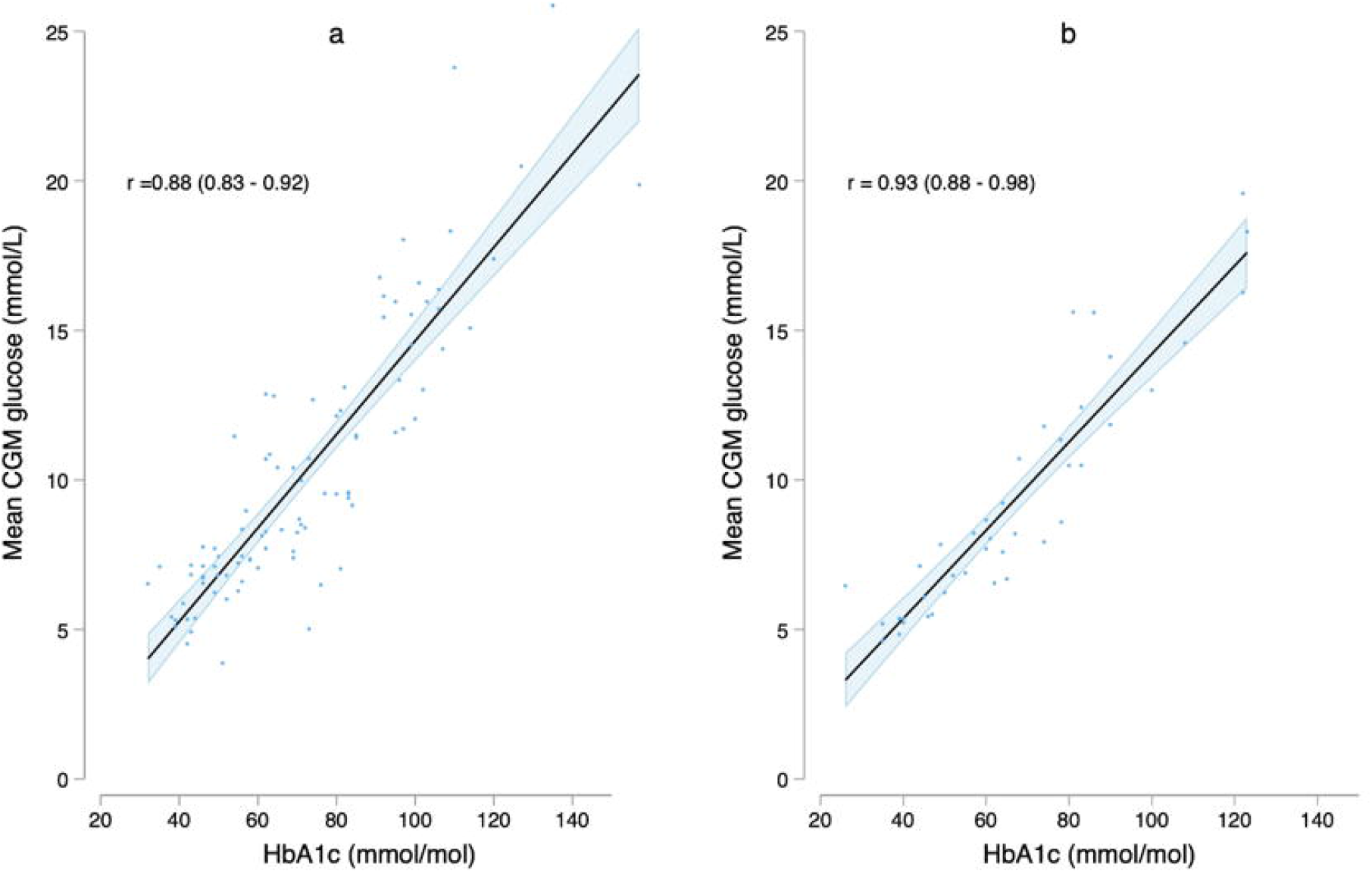
Correlation between HbA1c and CGM glucose. Scatterplots of the relationship between mean glucose obtained through continuous glucose monitoring and HbA1c in type 2 diabetes patients. Correlation between mean CGM and HbA1c in iron replete/normal iron state (a) and iron deplete state but without anemia (b). Solid straight line denotes the line of best fit and the side lines represent the 95% confidence interval. The Pearson’s correlation coefficient (r) and 95% confidence intervals are shown for each graph.

### Correlation between HbA1c with serum ferritin in both iron deplete and replete participants

The relationship between HbA1c and serum ferritin showed a weak positive correlation as indicated by the median regression line (Scatter plot). However, substantial data point dispersion around this regression line indicated high variability (S4 graph).

### Effect of iron deficiency on HbA1c levels

From the multivariable model (Table 2), iron deficiency had no significant effect on HbA1c levels (P=0.082). Hypertension and prolonged diabetes duration was associated with lower (−1.42%, 95% CI; −2.33 to −0.51, P=0.003) and higher (0.15%, 95% CI; 0.07 to 0.24, P<0.001) HbA1c levels respectively. Age, sex and BMI were not associated with significant changes in HbA1c levels.

**Table 2.**
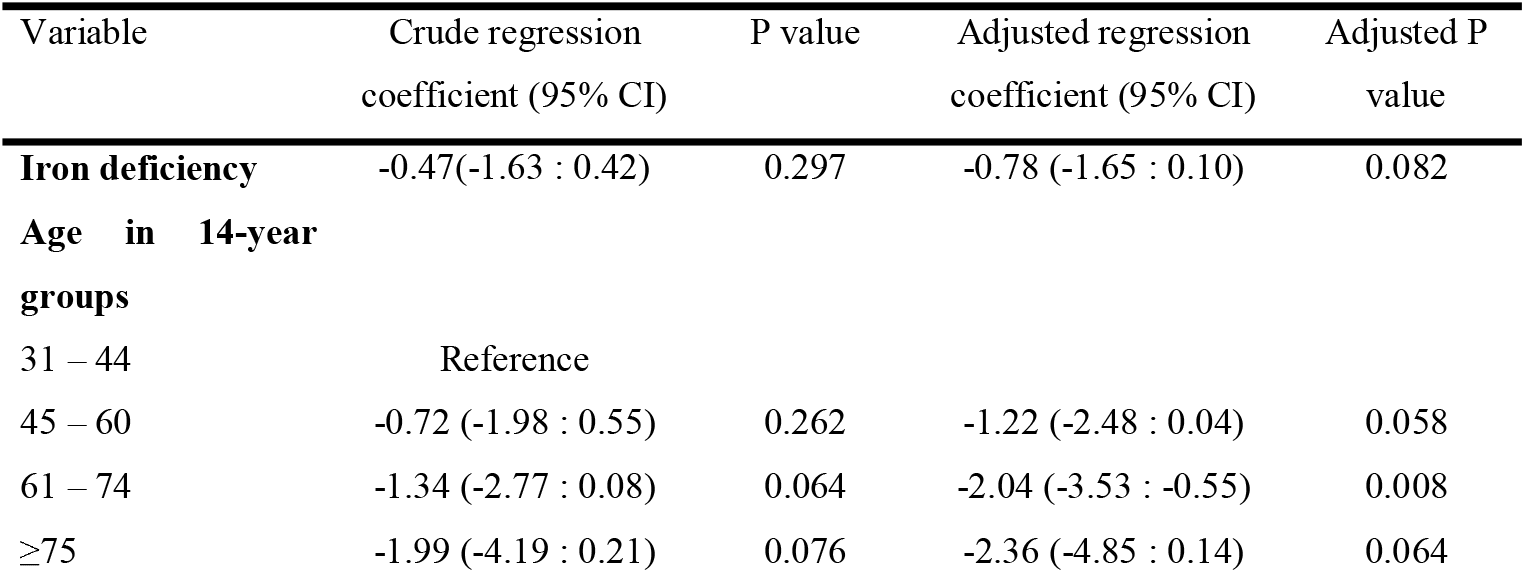
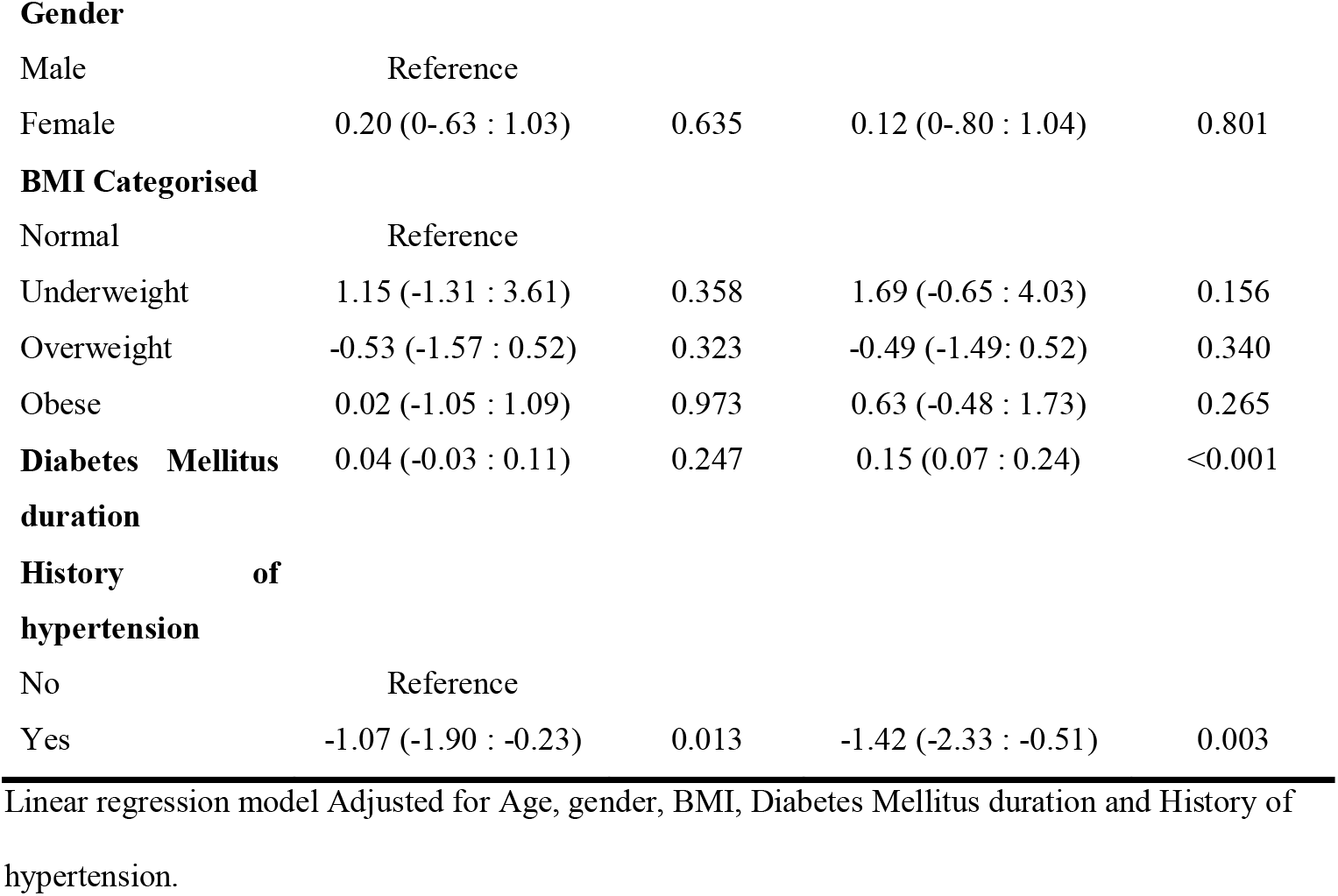
A linear regression model assessing the effect of the exposure (iron deficiency) against the outcome (HbA1c levels) adjusting for other a priori confounders.

## Discussion

In this study involving non-anaemic participants with Type 2 diabetes mellitus (T2DM), we found no significant differences in mean HbA1c concentrations between iron-replete and iron-deplete individuals and this result did not differ after adjusting for potential confounders i.e. age, gender, BMI, hypertension status and diabetes duration. Furthermore, we observed a strong positive correlation between HbA1c and mean CGM glucose in both groups, indicating that HbA1c is a dependable marker for monitoring glycaemic control in non-anaemic individuals with iron deficiency.

Similar results have been observed elsewhere by Heyningen et al. (16), Ford et al. (17) and Rai et al. (18) reporting no significant differences in HbA1c levels between iron-replete and iron-deficient individuals. Likewise, the strong positive correlation observed between HbA1c and mean plasma glucose levels in our study aligns with findings from previous research, which also reported a similar correlation between these two variables (19-21) However, these did not use continuous glucose monitoring (CGM) but used other measure of glucose burden for example fasting glucose to determine the correlation between mean plasma glucose and HBA1c. In another study, Hashimoto and Koga (22) observed a similar result among non-pregnant women, however, HbA1c levels significantly differed among the pregnant women. Other studies have reported contrasting results that iron deficiency impacts HbA1c levels amongst individuals. (23). Most of these studies however included methodological limitations, including small sample sizes and inadequate control for confounding factors such as anaemia and hemoglobinopathies

Our finding suggesting that iron deficiency does not significantly impact HbA1c levels in non-anaemic type 2 diabetes patients deserves further exploration to better understand the interaction between iron deficiency and HbA1c. One possible explanation is that iron, a crucial precursor in erythropoiesis (24), plays a key role in the formation of new RBCs. In iron deficiency, the reduced availability of iron may limit the production of new RBCs, leading to a higher proportion of older RBCs, which have been more exposed to glycaemia. This could result in the offsetting effect of older red blood cells on the younger, less glycation-prone RBCs. Furthermore, since HbA1c is influenced by RBC age, the reduced rate of erythropoiesis in iron-deficient patients may contribute to the observed stability of HbA1c measurements. This may explain why iron replacement therapy (potentially leading to the restoration of the erythropoietic process) in these patients is associated with reduction in HbA1c levels (as the production of new red blood cells normalizes)(25)

This study has clinical implications given the global burden of iron deficiency(26). Unlike anaemia, which can be easily screened, the diagnosis of iron deficiency is often more challenging requiring complex methods that are often unavailable in peripheral healthcare settings(27). For non-anaemic patients, our findings imply that there is no immediate clinical concern regarding iron status, and HbA1c can still be a reliable marker for glycaemic control. However, the present study has some limitations that should be taken into consideration. First, iron status in our study was assessed using serum ferritin, which is one of the most widely used markers of iron stores. However, it is important to note that serum ferritin levels can be influenced by inflammation (28, 29), a common condition in diabetes, potentially leading to inaccurate assessment of iron status. Given this limitation, future studies should consider using more reliable biomarkers such as soluble transferrin receptor (sTfR), which may provide a more accurate reflection of iron status independent of inflammation(30). We did not investigate the causes of iron deficiency in this study and therefore we were unable to do further subgroup analyses to assess the impact of the different underlying causes of iron deficiency (e.g., chronic blood loss versus decreased iron absorption)(27), so our results may differ between the different causes of iron deficiency. HbA1c reflects glycaemia over 90-120 days, a duration that is significantly longer than the duration of the current 14-day study period (31). Although we excluded participants with anaemia, renal impairment, and haemoglobinopathies, comorbidities that are known to affect HbA1c, we did not screen for other factors known to affect RBC e.g., glucose-6 phosphate dehydrogenase (G6PD) variants (32).

However, it should be noted that there is still limited data on how iron deficiency in the absence of anaemia impacts HbA1c as most of the conflicting results from previous studies were rather focussed more on assessing the impact of iron deficiency anaemia on HbA1c (17, 33-36).

In conclusion, HbA1c strongly correlates with mean blood glucose levels, even in iron-depleted states, among non-anaemic individuals with T2DM, supporting its reliability as a marker for glycemic control. However, larger studies are needed to confirm these findings, particularly at screening and diagnostic thresholds.

## Supporting information

Supplemental graph 1

Supplemental graph 2

Supplemental graph 3

Supplemental graph 4

## Authors’ contributions

## Conceptualisation

Moffat J Nyirenda, Anxious Jackson Niwaha.

## Data analysis

Isaac Ssekitoleko, Ronald Makanga, Terry Ongaria.

## Writing – original draft

Rogers John Mukasa, Anxious Jackson Niwaha, Isaac Ssekitoleko, Ronald Makanga.

## Writing – review & editing

Moffat J Nyirenda, Anxious Jackson Niwaha, Rogers John Mukasa, Isaac Ssekitoleko, Ronald Makanga, Nathan Mubiru, Hubert Nkabura, Terry Ongaria, Viola Mugamba, Wisdom Nakanga.

## Data availability

All the relevant data are within the paper and its Supporting information files.

## Acknowledgements

The authors are grateful to Aisha Karungi and Winnie Ayuru, the research assistants that carried out the study activities at the diabetes clinics, to the staff at the two laboratories CDLS laboratory (Uganda) and Exeter clinical laboratory (UK) that performed the various assays. We are thankful to the hospital ethical committees and staff of the diabetes clinics of Masaka regional referral hospital and St. Francis hospital, Nsambya in Uganda.

## Supplementary information

**S1 graph: Prevalence of iron deficiency (ID) by (a) site and by (b) site and sex (grey; female and white; male)**.

**S2 graph: Density distribution across five ferritin categories (<58, 58-90, 91-137, 138-246**, ≥**247** μ**g/L)**

**S3 graph: Correlation between HbA1c (mmol/mmol) and mean continuous glucose among iron-deplete cases (b) and their matched iron-replete controls (a)**.

Solid straight line denotes the line of best fit and the side lines represent the 95% confidence interval. The Pearson’s correlation coefficient (r) and 95% confidence intervals are shown for each graph.

**S4 graph: Correlation between HbA1c with serum ferritin in both iron deplete and replete participants**

## References

1. Thomas R, Halim S, Gurudas S, Sivaprasad S, Owens D. IDF Diabetes Atlas: A review of studies utilising retinal photography on the global prevalence of diabetes related retinopathy between 2015 and 2018. Diabetes research and clinical practice. 2019;157:107840.

2. Dunachie S, Chamnan P. The double burden of diabetes and global infection in low and middle-income countries. Transactions of the Royal Society of Tropical Medicine and Hygiene. 2019;113(2):56–64.

3. Standards of Medical Care in Diabetes–2006. Diabetes Care. 2006;29(Suppl 1):s4–s42.

4. Nitin S. HbA1c and factors other than diabetes mellitus affecting it. Singapore Med J. 2010;51(8):616–22.

5. Bhattacharjee R, Thukral A, Chakraborty PP, Roy A, Goswami S, Ghosh S, et al. Effects of thyroid status on glycated hemoglobin. Indian journal of endocrinology and metabolism. 2017;21(1):26.

6. Camaschella C. New insights into iron deficiency and iron deficiency anemia. Blood reviews. 2017;31(4):225–33.

7. Brooks A, Metcalfe J, Day J, Edwards M. Iron deficiency and glycosylated haemoglobin A1. The Lancet. 1980;316(8186):141.

8. Sluiter W, Van Essen L, Reitsma W, Doorenbos H. Glycosylated haemoglobin and iron deficiency. The Lancet. 1980;316(8193):531–2.

9. Ahmad J, Rafat D. HbA1c and iron deficiency: a review. Diabetes & Metabolic Syndrome: Clinical Research & Reviews. 2013;7(2):118–22.

10. Niwaha AJ, Rodgers LR, Greiner R, Balungi PA, Mwebaze R, McDonald TJ, et al. HbA1c performs well in monitoring glucose control even in populations with high prevalence of medical conditions that may alter its reliability: the OPTIMAL observational multicenter study. BMJ Open Diabetes Research and Care. 2021;9(1):e002350.

11. Davidson MB, Schriger DL. Effect of age and race/ethnicity on HbA1c levels in people without known diabetes mellitus: implications for the diagnosis of diabetes. Diabetes research and clinical practice. 2010;87(3):415–21.

12. Dubowitz N, Xue W, Long Q, Ownby J, Olson D, Barb D, et al. Aging is associated with increased HbA1c levels, independently of glucose levels and insulin resistance, and also with decreased HbA1c diagnostic specificity. Diabetic Medicine. 2014;31(8):927–35.

13. Hovestadt I, Kiess W, Lewien C, Willenberg A, Poulain T, Meigen C, et al. HbA1c percentiles and the association between BMI, age, gender, puberty, and HbA1c levels in healthy German children and adolescents. Pediatric diabetes. 2022;23(2):194–202.

14. Huang X, Qin C, Guo X, Cao F, Tang C. Association of hemoglobin A1c with the incidence of hypertension: A large prospective study. Frontiers in Endocrinology. 2023;13:1098012.

15. Verma M, Paneri S, Badi P, Raman P. Effect of increasing duration of diabetes mellitus type 2 on glycated hemoglobin and insulin sensitivity. Indian Journal of clinical biochemistry. 2006;21:142–6.

16. Heyningen Cv, Dalton R. Glycosylated haemoglobin in iron-deficiency anaemia. 1985.

17. Ford ES, Cowie CC, Li C, Handelsman Y, Bloomgarden ZT. Iron deficiency anemia, nonLironLdeficiency anemia and HbA1c among adults in the US. Journal of diabetes. 2011;3(1):67–73.

18. Rai K, Pattabiraman T. Glycosylated haemoglobin levels in iron deficiency anaemia. 1986.

19. Ito C, Maeda R, Ishida S, Sasaki H, Harada H. Correlation among fasting plasma glucose, two-hour plasma glucose levels in OGTT and HbA1c. Diabetes research and clinical practice. 2000;50(3):225–30.

20. Kilpatrick ES, Rigby AS, Atkin SL. Variability in the relationship between mean plasma glucose and HbA1c: implications for the assessment of glycemic control. Clinical chemistry. 2007;53(5):897–901.

21. Rohlfing CL, Wiedmeyer H-M, Little RR, England JD, Tennill A, Goldstein DE. Defining the relationship between plasma glucose and HbA1c: analysis of glucose profiles and HbA1c in the Diabetes Control and Complications Trial. Diabetes care. 2002;25(2):275–8.

22. Hashimoto K, Koga M. Influence of iron deficiency on HbA1c levels in pregnant women: comparison with non-pregnant women. Journal of Clinical Medicine. 2018;7(2):34.

23. Attard S, Herring A, Wang H, Howard A, Thompson A, Adair L, et al. Implications of iron deficiency/anemia on the classification of diabetes using HbA1c. Nutrition & diabetes. 2015;5(6):e166–e.

24. Abbaspour N, Hurrell R, Kelishadi R. Review on iron and its importance for human health. Journal of research in medical sciences: the official journal of Isfahan University of Medical Sciences. 2014;19(2):164.

25. AlQarni AM, Alghamdi AA, Aljubran HJ, Bamalan OA, Abuzaid AH, AlYahya MA. The effect of iron replacement therapy on HbA1c levels in diabetic and nondiabetic patients: a systematic review and meta-analysis. Journal of Clinical Medicine. 2023;12(23):7287.

26. Lopez A, Cacoub P, Macdougall IC, Peyrin-Biroulet L. Iron deficiency anaemia. The Lancet. 2016;387(10021):907–16.

27. Camaschella C. Iron-Deficiency Anemia. New England Journal of Medicine. 2015;372(19):1832–43.

28. Daru J, Colman K, Stanworth SJ, De La Salle B, Wood EM, Pasricha S-R. Serum ferritin as an indicator of iron status: what do we need to know?†‡§. The American Journal of Clinical Nutrition. 2017;106:1634S–9S.

29. Dignass A, Farrag K, Stein J. Limitations of Serum Ferritin in Diagnosing Iron Deficiency in Inflammatory Conditions. International Journal of Chronic Diseases. 2018;2018(1):9394060.

30. Rusch JA, van der Westhuizen DJ, Gill RS, Louw VJ. Diagnosing iron deficiency: Controversies and novel metrics. Best Practice & Research Clinical Anaesthesiology. 2023;37(4):451–67.

31. Tahara Y, Shima K. The response of GHb to stepwise plasma glucose change over time in diabetic patients. Diabetes Care. 1993;16(9):1313–4.

32. Wheeler E, Leong A, Liu CT, Hivert MF, Strawbridge RJ, Podmore C, et al. Impact of common genetic determinants of Hemoglobin A1c on type 2 diabetes risk and diagnosis in ancestrally diverse populations: A transethnic genome-wide meta-analysis. PLoS medicine. 2017;14(9):e1002383.

33. Firat A, Katlan DC, Uzunay N. Impact of Iron Deficiency Anemia on Hemoglobin A1c Levels in Diabetic and Non-Diabetic Pregnant Women. Clinical and Experimental Obstetrics & Gynecology. 2024;51(1):24.

34. Silva JF, Pimentel AL, Camargo JL. Effect of iron deficiency anaemia on HbA1c levels is dependent on the degree of anaemia. Clinical biochemistry. 2016;49(1-2):117–20.

35. Sinha N, Mishra T, Singh T, Gupta N. Effect of iron deficiency anemia on hemoglobin A1c levels. Annals of laboratory medicine. 2012;32(1):17.

36. GramLHansen P, Eriksen J, MouritsLAndersen T, Olesen L. Glycosylated haemoglobin (HbA1c) in iron and vitamin B12 deficiency. Journal of internal medicine. 1990;227(2):133–6.

