## Supplementary figures and images for "Impact of Iron Deficiency on HbA1c Accuracy in Monitoring Glycaemic Control in Non-Anaemic Type 2 Diabetes individuals: A prospective longitudinal study"

### Supplemental graph 1

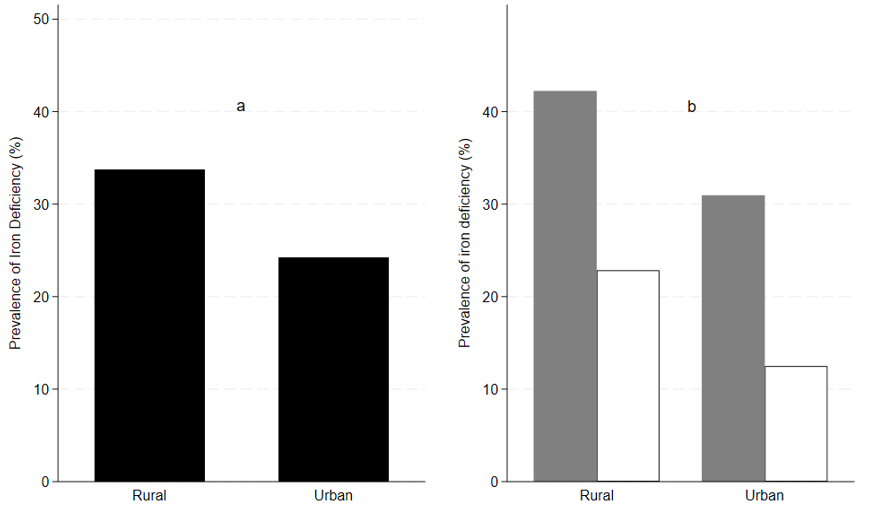

### Supplemental graph 2

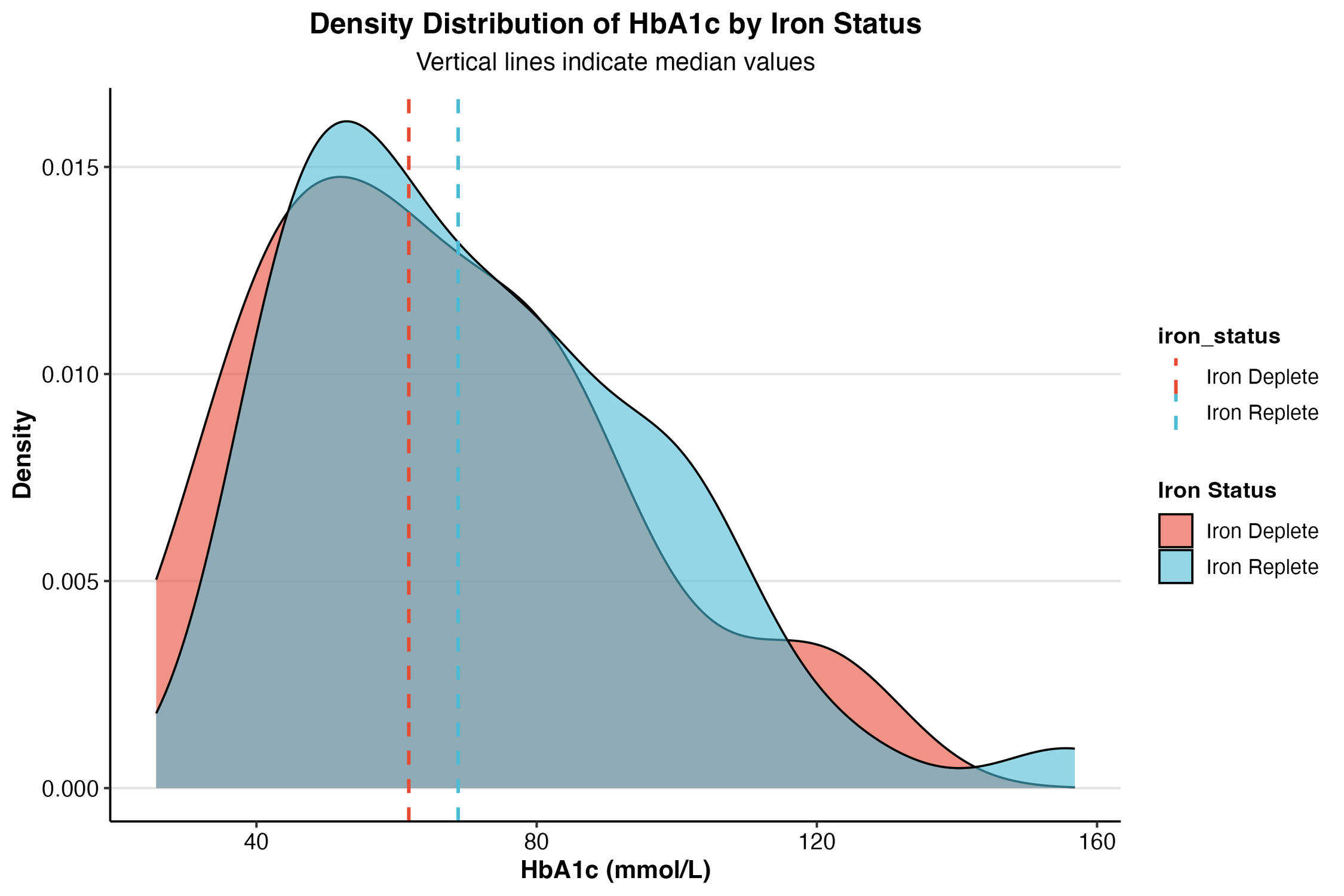

### Supplemental graph 3

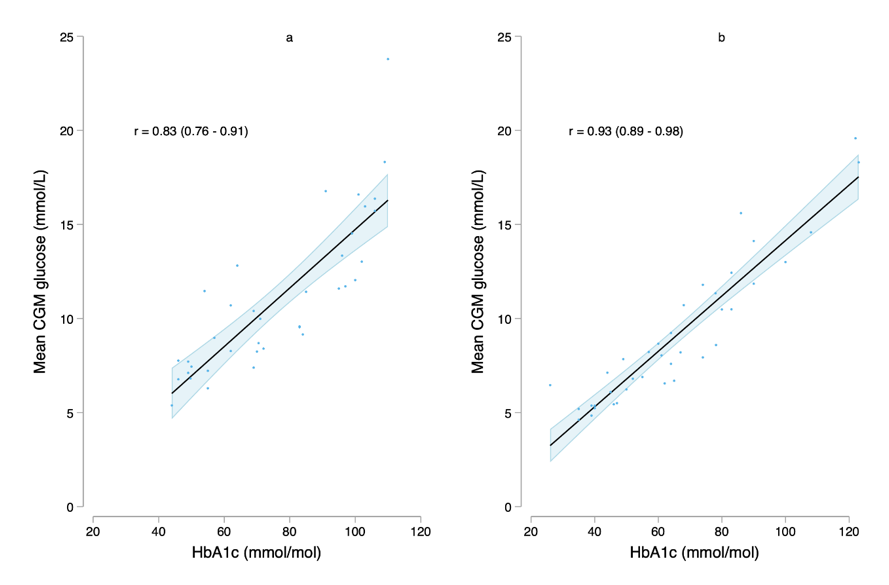

### Supplemental graph 4

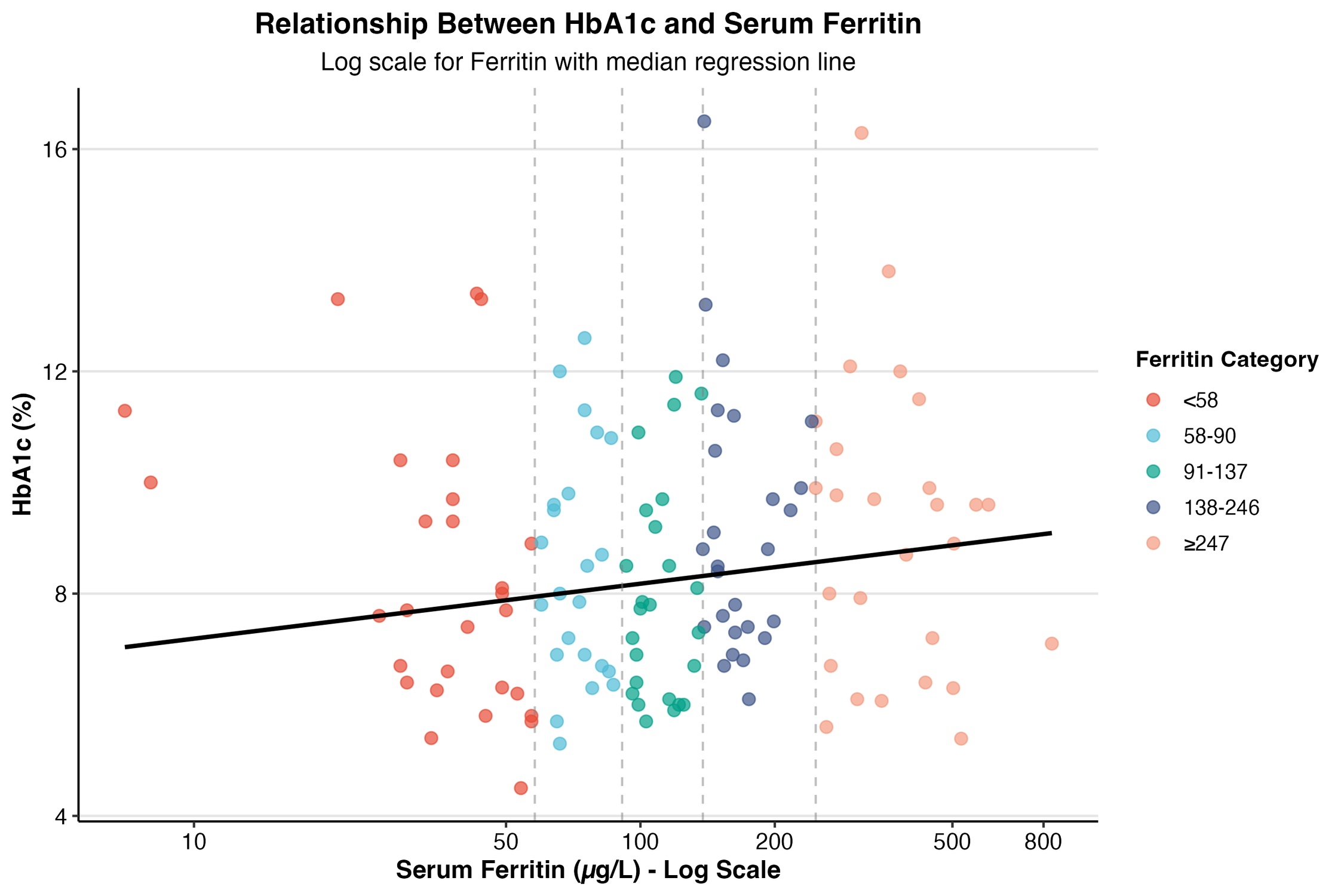
